# Ultra-processed food intake and risk of postmenopausal breast cancer in the NIH-AARP Diet and Health Study

**DOI:** 10.1101/2025.11.17.25340422

**Authors:** Caitlin P. O’Connell, Kaelyn F. Burns, Hyokyoung G. Hong, Lisa Kahle, Linda M. Liao, Jessica M. Madrigal, Camella J. Rising, Rashmi Sinha, Neha Khandpur, Eurídice Martínez Steele, Gretchen Gierach, Erikka Loftfield

## Abstract

**Background:** UPF intake is associated with obesity. Despite obesity being a risk factor for postmenopausal breast cancer, evidence for an association between UPF and breast cancer is limited. Our objective was to assess the association between UPF intake and postmenopausal breast cancer risk in the NIH-AARP Diet and Health Study.

**Methods:** Participants reported dietary intake via a food frequency questionnaire at baseline in 1995-1996 and were followed through 2018. Food items were disaggregated into food codes and assigned Nova classification via database linkage. Multivariable Cox proportional hazards regression models were used to estimate hazard ratios (HR) and 95% confidence intervals (CI) for quintiles of UPF (g/1000 kcal/day) and postmenopausal breast cancer risk overall, by estrogen receptor (ER) status, and type (invasive or ductal carcinoma in situ (DCIS)), with and without adjustment for body mass index (BMI).

**Results:** Among 181,460 postmenopausal women, 14,484 were diagnosed with breast cancer over a median 21 years of follow-up. Median (IQR) UPF intake was 284.6 (191.1-466.6) g/1000 kcal/day. No associations between UPF intake and postmenopausal breast cancer overall (HR_Q5vs.Q1_=0.98, 95% CI=0.93-1.03; P_trend_=0.32), by ER status (ER+: HR_Q5vs.Q1_=1.01, 95% CI=0.94-1.09; P_trend_=0.95; ER-: HR_Q5 vs. Q1_=1.02, 95% CI=0.87-1.20; P_trend_=0.58), or by type (invasive: HR_Q5vs.Q1_=0.98, 95% CI=0.92-1.04; P_trend_=0.14; DCIS HR_Q5vs.Q1_=1.00, 95% CI=0.87-1.13; P_trend_=0.26) were observed. Following BMI adjustment, inverse trends for overall (P_trend_=0.03) and invasive (P_trend_=0.01) breast cancer were observed.

**Conclusion:** UPF intake was not associated with postmenopausal breast cancer in the NIH-AARP cohort. Inverse trends observed after adjusting for BMI are likely spurious, owing to over-adjustment bias.

## INTRODUCTION

Breast cancer is the most commonly diagnosed cancer among women in the United States (US), accounting for an estimated 32% of new cancer cases in women in 2025.^1^ According to a 2024 report from the American Cancer Society, breast cancer incidence rates have increased by approximately 0.6% per year since the mid-2000s, driven primarily by hormone receptor-positive disease.^2,3^

Ultra-processed food (UPF) consumption has also increased in the US in recent decades^4^ and accounts for over half the caloric intake of the US population.^4,5^ High UPF intake has been associated with weight gain^6^ and obesity.^7-9^ Obesity is an established risk factor for postmenopausal breast cancer,^10^ particularly estrogen receptor (ER) positive cancer.^11,12^ However, the current literature is limited on the association of UPF with cancer.^7^ Furthermore, studies to-date that have investigated the association between UPF intake and risk of breast cancer, specifically, have found inconsistent results and have not considered ER status or breast cancer type (invasive, ductal carcinoma in situ (DCIS)).^13-15^ Using data from the NIH-AARP Diet and Health Study, the objective of our study was to assess the association between UPF intake and risk of postmenopausal breast cancer overall, by ER status, and by invasive and non-invasive type in a large US prospective cohort.

## METHODS

### Study population

The NIH-AARP Diet and Health Study began in 1995-1996 when approximately 3.5 million members of AARP, aged 50-69 years, residing in six states (Florida, Louisiana, New Jersey, North Carolina, Pennsylvania, and California) and two metropolitan areas (Atlanta, Georgia and Detroit, Michigan), were mailed baseline questionnaires querying demographic and lifestyle characteristics, including dietary habits.^16^ The NIH-AARP study was approved by the Special Studies Institutional Review Board of the US National Cancer Institute; informed consent was implied via return of a completed questionnaire. In total, 566,398 participants returned a questionnaire with satisfactory dietary data^16^ and were included in the cohort. We excluded those who had a proxy respondent (n=15,760), were male (n=325,171), had self-reported or registry-confirmed cancer at baseline (n=23,922), self-reported end-stage renal disease at baseline (n=371), had a death record for cancer but no registry confirmed diagnosis (n=5,251), had extreme calorie consumption defined as >2 interquartile ranges (IQR) below the 25^th^ or above the 75^th^ percentile of energy intake (n=1706), had <1 year of follow-up (n=3,809), or were premenopausal (i.e., self-reported “still menstruating”) or had unknown menopausal status (n=8,948) at baseline. Our final analytic cohort included 181,460 postmenopausal women.

### Exposure assessment

Participants in the NIH-AARP Diet and Health Study reported dietary intake via a food frequency questionnaire (FFQ) that asked about the intake of 124 food and beverage items over the past 12 months at baseline. Also included were 21 questions regarding intake of low-fat, high-fiber foods and food preparation.^16^ This resulted in 204 items for Nova classification. As previously described,^17^ FFQ line items were translated into individual food codes and further disaggregated into food and standard references codes via database linkage for the Nova classification approach described by MartÍnez Steele et al.^5^ Grams and energy values were then summed for each Nova group and subgroup for each participant. Our prior work indicated that UPF intake defined as absolute grams versus percentage grams or percentage energy was the measure with highest validity^17^; therefore, quintiles of nutrient-density adjusted^18^ grams of UPF (g/1000 kcal/day) were our main exposure variable. We excluded alcoholic beverages from our Nova definition as we decided a priori to adjust for alcohol intake, an established risk factor for breast cancer.^19,20^

### Outcome ascertainment

Cancer cases were ascertained through probabilistic linkages to the eight states of baseline residency and three popular relocation states (Arizona, Texas, and Nevada).^21^ Vital status was determined via linkage to the National Death Index. Incident first primary breast cancer cases were identified using the SEER cancer site recode of the International Classification of Disease for Oncology, Third Edition (ICD-O-3).^22^ Breast cancer cases were additionally classified as invasive or DCIS, defined with the following histologies: 8201, 8230, 8500, 8501, 8503, 8507, 8523. We also classified breast cancer cases by ER status (e.g., ER+ or ER-) regardless of progesterone receptor (PR) status) since only 1% of breast cancers with data on hormone receptor status were ER-/PR+ (n=112). Participants who were missing information on ER status were censored at date of diagnosis (31.3%; n=4539). Follow-up time in years was defined as baseline to date of any cancer diagnosis, death, relocation out of recruitment area, or end of study follow-up (December 31, 2018). Exit age in years was calculated based on entry age at baseline and follow-up time.

### Statistical analysis

We tabulated demographic and lifestyle factors by quintile of nutrient-density adjusted grams of UPF per 1000 kcal per day. We used Cox proportional hazards regression models to estimate hazard ratios (HR) and 95% confidence intervals (CI) for the association of UPF intake and risk of breast cancer, overall, by type (invasive vs. DCIS), and by ER status (ER+ vs. ER-; regardless of PR status). Our first model was adjusted for age (underlying time metric) and energy (kcal/day). Our second model was further adjusted for the following potential confounders: race and ethnicity (American Indian/Alaskan Native, Asian, Hispanic, non-Hispanic Black, non-Hispanic White, Pacific Islander, or unknown), smoking status, incorporating intensity and time since cessation (categorical), education (<12 years; 12 years, completed high school, GED; post-high school training; some college; college and post graduate; unknown), physical activity (never, 1-3 times/month, 1-2 times/week, ≥3 times/week, unknown), alcohol intake (0 drinks per day, <1 drink per day, 1-2 drinks per day, 3-4 drinks per day, ≥5 drinks per day), marital status (married, widowed, divorced or separated, never married, unknown), years of hormone replacement therapy (HRT) (never, <5, 5-9, ≥10 years, unknown), age at first menstrual period (≤10, 11-12, 13-14, ≥15 years, unknown), years of oral contraceptive use (never or <1 year, 1-4 years, ≥5 years, unknown), parity (none, 1, ≥2, unknown), and first degree relative with breast cancer (yes, no, unknown). To assess whether the association was independent of BMI status, we ran a third model further adjusting for BMI category. An indicator variable was used for covariates with missing data; no covariate had more than 4.5% of data missing. To assess a linear trend across UPF quintiles, we assigned each quintile to its median intake and treated this as a continuous variable. The proportional hazards assumption was evaluated using the global score test and by inspecting Schoenfeld residual plots.^23-25^ The proportional hazards assumption was not violated (global score test p > 0.05), and inspection of Schoenfeld residual plots revealed no evidence of time-dependent trends in the covariate effects. We further conducted a lag analysis, estimating HR estimates for UPF intake (g/1000 kcal/day) and breast cancer risk during 5-year follow-up periods.

In substitution and addition models, we evaluated specific dietary factors (e.g., calcium and saturated fat) and food groups (e.g., vegetables and total meat) that have been previously associated with breast cancer risk.^26-28^ In substitution models, we included total dietary intake (Nova groups 1-4, g/1000 kcal/day) and total dietary intake from UPF sources (Nova group 4) in the models, such that an increase in intake from UPF is offset by an equal decrease in intake from non-UPF sources while holding total intake constant.^29,30^ For example, total dietary calcium and dietary calcium from UPF sources were included in the calcium substitution model, such that an increase in dietary calcium from UPF (Nova group 4) sources is offset by an equal decrease in dietary calcium from non-UPF (Nova groups 1-3) sources while holding total dietary calcium (from UPF and non-UPF) intake constant. In addition models, we mutually adjusted for UPF intake and non-UPF intake of dietary calcium, saturated fat, vegetables, and meat such that UPF intake of dietary calcium is independent of non-UPF intake of dietary calcium and other adjustment variables.

We also reran our main models for postmenopausal breast cancer using quintiles of UPF based on percentage grams per day and percentage energy per day.

All analyses were conducted using RStudio, version 4.3.3.^31^ A two-sided P-value <.05 was considered statistically significant.

## RESULTS

Overall, median intake of UPF was 284.6 g/1000 kcal/day (IQR=191.1-466.6) and accounted for 42.2% of total energy intake (IQR=35.3-49.2). Women in the highest quintile (Q5) of UPF intake (median [IQR]: 756.0 [617.6-1026.9] g/1000 kcal/day), compared to the lowest quintile (Q1; 146.8 [126.4-162.5]), were younger (60.9 vs. 63.9 years old), and were more likely to be non-Hispanic black (6.3% vs. 5.1%) or non-Hispanic white (90.3% vs. 86.9%), current smokers (17.5% vs. 15.1%), never exercise (27.3% vs. 20.4%), nondrinkers (36.0% vs. 28.4%), obese (31.1% vs. 15.4%), or married (43.6% vs. 41.9%) (Table 1; Supplementary table 1). Those with the highest compared to lowest UPF intake were less likely to have graduated from college (25.2% vs. 32.2%), used HRT (44.6% vs. 46.2%), start their menstrual period ≥15 years old (9.0% vs. 10.5%), use oral contraceptives (57.0% vs. 64.4%), be nulliparous (14.1% vs. 16.3%) or have a first degree relative with breast cancer (11.9% vs. 12.4%).

**Table 1.** Baseline characteristics by nutrient-adjusted ultra-processed food intake ^a^ in NIH-AARP breast cancer analytic cohort (N=181,460)

| Characteristic, N (%) | Quintile 1<br>(N=36292) | Quintile 2<br>(N=36292) | Quintile 3<br>(N=36292) | Quintile 4<br>(N=36292) | Quintile 5<br>(N=36292) |
| --- | --- | --- | --- | --- | --- |
| Median UPF intake, g/1000 kcal/day [IQR] | 146.8 [126.4, 162.5] | 206.1 [191.1, 222.7] | 284.6 [261.0, 311.8] | 417.3 [377.4, 466.6] | 756.0 [617.6, 1026.9] |
| Median UPF intake, % kcal/day [IQR] | 34.0 [28.0, 40.1] | 41.5 [35.7, 47.7] | 43.3 [37.4, 49.6] | 44.8 [38.7, 51.1] | 47.2 [40.0, 54.8] |
| Median UPF intake, % g/day [IQR] | 8.6 [6.5, 11.2] | 13.1 [10.5, 16.6] | 18.1 [14.6, 22.7] | 25.0 [20.4, 31.0] | 40.3 [31.5, 51.9] |
| Median age at baseline, year [IQR] | 63.9 [59.2, 67.4] | 63.4 [58.8, 67.0] | 62.8 [58.2, 66.6] | 62.1 [57.7, 66.1] | 60.9 [56.7, 65.3] |
| Race/ethnicity |  |  |  |  |  |
| American Indian/Alaskan Native | 130 (0.4%) | 85 (0.2%) | 118 (0.3%) | 112 (0.3%) | 154 (0.4%) |
| Asian | 1038 (2.9%) | 373 (1.0%) | 294 (0.8%) | 190 (0.5%) | 123 (0.3%) |
| Hispanic | 993 (2.7%) | 805 (2.2%) | 693 (1.9%) | 576 (1.6%) | 426 (1.2%) |
| Non-Hispanic Black | 1846 (5.1%) | 1754 (4.8%) | 2055 (5.7%) | 2226 (6.1%) | 2283 (6.3%) |
| Non-Hispanic White | 31542 (86.9%) | 32695 (90.1%) | 32615 (89.9%) | 32661 (90.0%) | 32773 (90.3%) |
| Pacific Islander | 88 (0.2%) | 41 (0.1%) | 47 (0.1%) | 28 (0.1%) | 22 (0.1%) |
| Unknown | 655 (1.8%) | 539 (1.5%) | 470 (1.3%) | 499 (1.4%) | 511 (1.4%) |
| Current smoker | 5462 (15.1%) | 4546 (12.5%) | 4320 (11.9%) | 4832 (13.3%) | 6339 (17.5%) |
| College graduate | 11669 (32.2%) | 11504 (31.7%) | 10783 (29.7%) | 10021 (27.6%) | 9151 (25.2%) |
| Never or rarely exercise | 7402 (20.4%) | 7320 (20.2%) | 7604 (21.0%) | 8366 (23.1%) | 9910 (27.3%) |
| 0 alcoholic drinks per day | 10322 (28.4%) | 9547 (26.3%) | 9982 (27.5%) | 10876 (30.0%) | 13074 (36.0%) |
| Body mass index $\geq 30$ kg/m<sup>2</sup> | 5575 (15.4%) | 6621 (18.2%) | 7905 (21.8%) | 9284 (25.6%) | 11288 (31.1%) |
| Married | 15195 (41.9%) | 16014 (44.1%) | 16302 (44.9%) | 16759 (46.2%) | 15821 (43.6%) |
| Never used hormone replacement therapy | 16760 (46.2%) | 15512 (42.7%) | 15487 (42.7%) | 15712 (43.3%) | 16190 (44.6%) |
| Age at first period, $\geq 15$ years | 3805 (10.5%) | 3368 (9.3%) | 3344 (9.2%) | 3248 (8.9%) | 3282 (9.0%) |
| Oral contraceptive use, never or < 1 yr | 23372 (64.4%) | 22625 (62.3%) | 22043 (60.7%) | 21356 (58.8%) | 20676 (57.0%) |
| Parity, none | 5908 (16.3%) | 5563 (15.3%) | 5354 (14.8%) | 4993 (13.8%) | 5103 (14.1%) |
| First degree relative with breast cancer | 4491 (12.4%) | 4557 (12.6%) | 4606 (12.7%) | 4470 (12.3%) | 4303 (11.9%) |

Abbreviations: UPF, Ultra-processed food; IQR, interquartile range; BMI, body mass index; HRT, hormone replacement therapy
<sup>a</sup> Quintiles of energy density-adjusted gram weight UPF (i.e., Nova group 4) intake (g/1000 kcal/day)

Over a median of 21 years of follow-up, 14,484 postmenopausal women were diagnosed with breast cancer. Of these, 11,854 were classified as invasive while 2,287 were classified as DCIS, and 8,366 were ER+ breast cancer while 1,579 were ER negative. We observed no association between UPF intake and risk of postmenopausal breast cancer (HR_Q5 vs. Q1_=0.98, 95% CI=0.93-1.03; P_trend_=0.32; Table 2). We also observed no association for risk of invasive breast cancer (HR=0.98, 0.92-1.04; P_trend_=0.14) or DCIS (HR=1.00, 0.87-1.13; P_trend_=0.26). Additionally, we observed no association for breast cancer risk defined by ER status (HR_ER+_=1.01, 0.96-1.09; P_trend_=0.95; HR_ER-_=1.02, 0.87-1.20; P_trend_=0.58). Adjustment for BMI status category slightly shifted the inverse HR estimate for breast cancer overall away from the null (HR=0.95, 0.90-1.00; P_trend_=0.03) as well as the inverse HR estimate for invasive breast cancer (HR=0.95, 0.98-1.00; P_trend_=0.008).

**Table 2.** Association of quintiles of ultra-processed food intake (g/1000 kcal/day) with breast cancer risk in the NIH-AARP Diet and Health Study (N=181,460)

| Cancer type | N | Q1 HR (95% CI)<br>(N=36292) | Q2 HR (95% CI)<br>(N=36292) | Q3 HR (95% CI)<br>(N=36292) | Q4 HR (95% CI)<br>(N=36292) | Q5 HR (95% CI)<br>(N=36292) | P-trend<sup>a</sup> |
| --- | --- | --- | --- | --- | --- | --- | --- |
| Breast cancer |  |  |  |  |  |  |  |
| No. of cases | 14484 | 2920 | 2931 | 3010 | 2845 | 2778 |  |
| Minimally adjusted model<sup>b</sup> |  | 1.00 (ref) | 0.99 (0.94-1.05) | 1.02 (0.97-1.07) | 0.97 (0.92-1.02) | 0.96 (0.91-1.01) | .05 |
| Multivariable adjusted model<sup>c</sup> |  | 1.00 (ref) | 0.99 (0.94-1.04) | 1.02 (0.97-1.07) | 0.98 (0.93-1.03) | 0.98 (0.93-1.03) | .32 |
| Multivariable adjusted + BMI model<sup>d</sup> |  | 1.00 (ref) | 0.98 (0.93-1.04) | 1.01 (0.96-1.06) | 0.96 (0.91-1.01) | 0.95 (0.90-1.00) | .03 |
| ER positive breast cancer |  |  |  |  |  |  |  |
| No. of cases | 8366 | 1678 | 1726 | 1746 | 1646 | 1570 |  |
| Minimally adjusted model<sup>b</sup> |  | 1.00 (ref) | 1.02 (0.96-1.10) | 1.04 (0.97-1.11) | 1.00 (0.93-1.07) | 0.99 (0.92-1.06) | .33 |
| Multivariable adjusted model<sup>c</sup> |  | 1.00 (ref) | 1.02 (0.95-1.09) | 1.04 (0.97-1.12) | 1.01 (0.94-1.08) | 1.01 (0.94-1.09) | .95 |
| Multivariable adjusted + BMI model<sup>d</sup> |  | 1.00 (ref) | 1.01 (0.94-1.08) | 1.02 (0.96-1.10) | 0.98 (0.91-1.05) | 0.97 (0.90-1.04) | .18 |
| ER negative breast cancer |  |  |  |  |  |  |  |
| No. of cases | 1579 | 315 | 302 | 329 | 314 | 319 |  |
| Minimally adjusted model<sup>b</sup> |  | 1.00 (ref) | 0.95 (0.81-1.11) | 1.03 (0.89-1.21) | 1.00 (0.85-1.16) | 1.04 (0.89-1.22) | .45 |
| Multivariable adjusted model<sup>c</sup> |  | 1.00 (ref) | 0.94 (0.80-1.10) | 1.01 (0.87-1.18) | 0.97 (0.83-1.14) | 1.02 (0.87-1.20) | .58 |
| Multivariable adjusted + BMI model<sup>d</sup> |  | 1.00 (ref) | 0.94 (0.80-1.10) | 1.02 (0.87-1.19) | 0.98 (0.84-1.15) | 1.03 (0.88-1.21) | .46 |
| Invasive breast cancer |  |  |  |  |  |  |  |
| No. of cases | 11854 | 2373 | 2448 | 2475 | 2309 | 2249 |  |
| Minimally adjusted model<sup>b</sup> |  | 1.00 (ref) | 1.02 (0.97-1.08) | 1.03 (0.97-1.09) | 0.97 (0.91-1.02) | 0.96 (0.91-1.02) | .03 |
| Multivariable adjusted model<sup>c</sup> |  | 1.00 (ref) | 1.02 (0.96-1.08) | 1.03 (0.98-1.10) | 0.98 (0.92-1.04) | 0.98 (0.92-1.04) | .14 |
| Multivariable adjusted + BMI model<sup>d</sup> |  | 1.00 (ref) | 1.01 (0.96-1.07) | 1.02 (0.96-1.08) | 0.96 (0.90-1.01) | 0.95 (0.89-1.00) | .01 |
| Ductal carcinoma in situ |  |  |  |  |  |  |  |
| No. of cases | 2287 | 477 | 409 | 470 | 469 | 462 |  |
| Minimally adjusted model<sup>b</sup> |  | 1.00 (ref) | 0.85 (0.74-0.97) | 0.97 (0.85-1.10) | 0.96 (0.85-1.10) | 0.96 (0.85-1.10) | .63 |
| Multivariable adjusted model<sup>c</sup> |  | 1.00 (ref) | 0.84 (0.74-0.96) | 0.96 (0.85-1.09) | 0.97 (0.86-1.11) | 1.00 (0.87-1.13) | .26 |
| Multivariable adjusted + BMI model<sup>d</sup> |  | 1.00 (ref) | 0.84 (0.73-0.96) | 0.95 (0.84-1.09) | 0.96 (0.85-1.10) | 0.98 (0.86-1.12) | .38 |

Abbreviations: HR, hazard ratio; CI, confidence interval; BMI, body mass index
<sup>a</sup>Each quintile was assigned to its median value and treated as a continuous variable.
<sup>b</sup>Adjusted for age in years (underlying time metric) and total daily energy (kcal/day) for nutrient density adjustment.
<sup>c</sup>Adjusted for age in years (underlying time metric), total daily energy (kcal/day), race/ethnicity (American Indian/Alaskan Native, Asian, Hispanic, Non-Hispanic Black, Non-Hispanic White, Pacific Islander, Unknown), smoking by intensity (cigarettes per day: 1-10, 11-20, 21-30, 31-40, 41-60, 61+) and time since cessation (10+ years ago, 5-9 years ago, 1-4 years ago, within the last year), education level (<12 years; 12 years, completed high school, or GED; post-high school training; some college; college and post graduate; unknown), physical activity level (never/rarely, 1-3 times/month, 1-2 times/week, ≥3 times/week, unknown), alcohol intake (0 drinks/day, < 1 drink/day, 1-2 drinks/day, 3-4 drinks/day, ≥5 drinks/day), first degree relative with breast cancer (yes/no), marital status (married, widowed, divorced or separated, never married, unknown), duration of HRT (never, <5, 5-9, ≥10 years, unknown), age at first menstrual period (<11, 11-12, 13-14, ≥15 years old, unknown), duration of oral contraceptive use (never or <1, 1-4, ≥5 years, unknown), and parity (0, 1, ≥2, unknown).
<sup>d</sup>Multivariable model further adjusted for body mass index category (<18.5 kg/m<sup>2</sup>, 18.5 to <25 kg/m<sup>2</sup>, 25 to <30 kg/m<sup>2</sup>, ≥30 kg/m<sup>2</sup>, unknown).

In our lag analysis, we observed an elevated, albeit not statistically significant, risk of breast cancer when follow-up was limited to the first five years (HR=1.08, 0.98-1.06; P_trend_=.21; Table 3). HR estimates were similar after the first five years and reflective of the overall estimate.

**Table 3.** Association of quintiles of ultra-processed food intake (g/1000 kcal/day) with breast cancer risk during 5-year follow up periods (1to <5 y, 5 to <10 y, 10 to ≤15 y, >15 y) in the NIH-AARP Diet and Health Study (N=181,460)

| Breast cancer | N | Q1 HR (95% CI) | Q2 HR (95% CI) | Q3 HR (95% CI) | Q4 HR (95% CI) | Q5 HR (95% CI) | P-trend <sup>a</sup> |
| --- | --- | --- | --- | --- | --- | --- | --- |
| Overall |  |  |  |  |  |  |  |
| No. of participants | 181460 | 36292 | 36292 | 36292 | 36292 | 36292 |  |
| No. of cases | 14484 | 2920 | 2931 | 3010 | 2845 | 2778 |  |
| Multivariable adjusted model <sup>b</sup> |  | 1.00 (ref) | 0.99 (0.94-1.04) | 1.02 (0.97-1.07) | 0.98 (0.93-1.03) | 0.98 (0.93-1.03) | .32 |
| 1 to <5 years |  |  |  |  |  |  |  |
| No. of participants | 181460 | 36292 | 36292 | 36292 | 36292 | 36292 |  |
| No. of cases | 3695 | 745 | 753 | 775 | 702 | 720 |  |
| Multivariable adjusted model <sup>b</sup> |  | 1.00 (ref) | 1.02 (0.92-1.13) | 1.08 (0.97-1.19) | 1.01 (0.91-1.12) | 1.08 (0.98-1.20) | .21 |
| 5 to <10 years |  |  |  |  |  |  |  |
| No. of participants | 164653 | 32776 | 32931 | 33009 | 33091 | 32846 |  |
| No. of cases | 4007 | 824 | 785 | 841 | 801 | 756 |  |
| Multivariable adjusted model <sup>b</sup> |  | 1.00 (ref) | 0.95 (0.86-1.05) | 1.03 (0.93-1.13) | 0.99 (0.89-1.09) | 0.96 (0.86-1.06) | .49 |
| 10 to ≤15 years |  |  |  |  |  |  |  |
| No. of participants | 141535 | 28034 | 28420 | 28502 | 28516 | 28063 |  |
| No. of cases | 3327 | 688 | 673 | 693 | 658 | 615 |  |
| Multivariable adjusted model <sup>b</sup> |  | 1.00 (ref) | 0.96 (0.86-1.07) | 0.99 (0.89-1.10) | 0.95 (0.86-1.06) | 0.92 (0.82-1.03) | .16 |
| >15 years |  |  |  |  |  |  |  |
| No. of participants | 120828 | 23872 | 24318 | 24485 | 24377 | 23776 |  |
| No. of cases | 3455 | 663 | 720 | 701 | 684 | 687 |  |
| Multivariable adjusted model <sup>b</sup> |  | 1.00 (ref) | 1.04 (0.94-1.16) | 0.99 (0.89-1.10) | 0.97 (0.87-1.08) | 0.99 (0.89-1.10) | .54 |

Abbreviations: HR, hazard ratio; CI, confidence interval; BMI, body mass index
<sup>a</sup>Each quintile was assigned to its median value and treated as a continuous variable.
<sup>c</sup>Adjusted for age in years (underlying time metric), total daily energy (kcal/day), race/ethnicity (American Indian/Alaskan Native, Asian, Hispanic, Non-Hispanic Black, Non-Hispanic White, Pacific Islander, Unknown), smoking by intensity (cigarettes per day: 1-10, 11-20, 21-30, 31-40, 41-60, 61+) and time since cessation (10+ years ago, 5-9 years ago, 1-4 years ago, within the last year), education level (<12 years; 12 years, completed high school, or GED; post-high school training; some college; college and post graduate; unknown), physical activity level (never/rarely, 1-3 times/month, 1-2 times/week, ≥3 times/week, unknown), alcohol intake (0 drinks/day, < 1 drink/day, 1-2 drinks/day, 3-4 drinks/day, ≥5 drinks/day), first degree relative with breast cancer (yes/no), marital status (married, widowed, divorced or separated, never married, unknown), duration of HRT (never, <5, 5-9, ≥10 years, unknown), age at first menstrual period (<11, 11-12, 13-14, ≥15 years old, unknown), duration of oral contraceptive use (never or <1, 1-4, ≥5 years, unknown), and parity (0, 1, ≥2, unknown).

Only 32.2%, 7.6%, and 15.2% of dietary calcium, vegetable, and meat intake was from UPF sources, respectively, while 45.9% of saturated fat was from UPF sources (Supplementary table 2). In the exploratory substitution analysis, substituting calcium from UPF sources for calcium from non-UPF sources was associated with a lower risk of breast cancer (HR=0.89, 0.82-0.97; Supplementary figure 1), while substitution of vegetables, saturated fat, or meat from UPF for intake from non-UPF was not associated with changes in breast cancer risk. In the addition model analysis, intake of calcium from UPF sources was associated with lower risk of breast cancer (HR=0.89, 0.82-0.96), while saturated fat from non-UPF sources was associated with higher risk of breast cancer (HR=1.08, 1.02-1.15). All other associations were null (Supplementary figure 1).

HR estimates were similar when UPF was operationalized as daily percentage grams or percentage energy (Supplementary tables 3 & 4).

## DISCUSSION

In a prospective cohort of 181,460 postmenopausal women with over 21 years of follow-up, UPF intake was not associated with risk of breast cancer overall, by ER status, or by type (invasive vs. DCIS). In contrast, we found intake of dietary calcium from UPF sources (32% of total dietary calcium) was associated with a lower risk of breast cancer.

Only three prospective cohort studies to-date have previously published on the relationship between UPF intake and risk of breast cancer. Our findings align with results of studies conducted in the UK Biobank and the EPIC cohort, which also observed no association.^14,15^ A study conducted in the NutriNet-Santé cohort reported an increased risk of postmenopausal breast cancer among 29,191 French women.^13^ While their assessment of UPF was conducted via multiple 24-hour recalls, a method arguably superior to our FFQ, the 24-hour recalls were collected across the first two years of the limited 5 years of follow-up. Therefore, participants could have reported intake after being diagnosed with breast cancer. Our study and two other cohort studies that similarly found null results^14,15^ had much longer follow-up periods of approximately 21,14, and 10 years, respectively, compared to 5 years in the NutriNet-Santé. In our lag analysis, we did observe an elevated risk of breast cancer, albeit not statistically significant, during the first five years of follow-up. As most carcinogenic processes may take decades to develop, extended follow-up time is warranted for this outcome.

It is plausible that higher intake of UPF is associated with breast cancer risk through weight gain and obesity since UPF has been linked to weight gain in two randomized controlled feeding trials,^6,32^ higher UPF intake has been associated with obesity in many prospective cohort studies,^9^ and adult body fatness is a risk factor for postmenopausal breast cancer,^10,26^ particularly ER+ cancers.^12^ However, the World Cancer Research Fund (WCRF) meta-analyzed relative risk estimate per 5 kg/m^2^ increase in BMI with postmenopausal breast cancer is relatively modest at RR=1.12 (95%CI=1.09-1.15).^26,33^ With a relatively small incremental increase in risk and diet not being the only determinant of BMI, a null association between higher UPF intake and risk of postmenopausal breast cancer may not be all that surprising, especially given the limited variation in intake between quintiles in our sample. Adjusting for BMI produced an inverse association for UPF intake with overall and invasive breast cancer. These findings should be interpreted with caution since BMI is modestly correlated with UPF intake in NIH-AARP and is a known risk factor for breast cancer, which suggests that BMI may be a mediator in the relationship between UPF intake and breast cancer. Adjusting for a mediator such as BMI can remove part of the pathway through which UPF intake may influence breast cancer risk and can introduce over adjustment bias.

Finally, there are few established dietary risk factors for breast cancer. Based on the WCRF 2018 Diet, Physical Activity and Cancer update report, the only convincing dietary risk factor for postmenopausal breast cancer is alcoholic drinks.^26^ While there is some evidence to suggest that higher intakes of non-starchy vegetables, foods containing carotenoids, and diets high in calcium decrease risk of postmenopausal breast cancer, the evidence is limited.^26^

### Strengths and limitations

Our study is not without limitations. Our exposure of UPF was measured by a FFQ, and measurement error is inherent to the FFQ. In addition, the FFQ was not designed to measure degree of food processing; however, our prior work demonstrated moderately high validity of our energy-adjusted grams measure of UPF.^17^ We would expect any measurement error in UPF intake to be non-differential by breast cancer status as the study is prospective in nature. Still, food group and nutrient analyses, which may be subject to a higher degree of measurement error, should be viewed as exploratory and interpreted with caution. Additionally, assessment of UPF was only conducted at baseline, over 20 years ago. Participants could have changed their intake over the course of follow-up. With trends of increasing consumption of UPF over the past two decades,^4^ our UPF estimates are most likely conservative in comparison to present day. Variation across quintiles of UPF intake was also limited, which may contribute to our null findings. Lastly, our cohort is comprised of older, predominantly non-Hispanic White, US adults, potentially limiting its generalizability to other populations. Studies combining cohorts from different countries and backgrounds should be conducted to confirm and strengthen the existing literature.

Despite these limitations, our study has notable strengths, including the large cohort size with sufficient statistical power to detect modest associations, long period of follow-up, and a Nova-defined UPF variable that has been validated using repeat 24-hour dietary recalls. To our knowledge, this is the first study to explore the association of UPF intake and breast cancer risk by ER status and invasive or non-invasive type.

## Conclusion

Our primary interpretation is based on the model without BMI because it more appropriately reflects the overall association between UPF intake and breast cancer in this cohort. We found no evidence of an association between higher UPF intake and postmenopausal breast cancer risk. This was consistent across ER+, ER-, invasive, and non-invasive tumors. Adjusting for BMI appears to have produced an inverse association for overall breast cancer that is likely the result of over adjustment and unlikely to reflect the true association between UPF intake and breast cancer. Further research with more diverse cohorts and detailed information on food processing should be conducted to confirm these findings.

## Data availability

Data are maintained by the National Cancer Institute, Division of Cancer Epidemiology and Genetics and are available upon approval of a proposal submitted to the NIH-AARP Diet and Health Study Steering Committee. For more information visit https://www.nihaarpstars.com/.

