## Supplemental material for "Ultra-processed food intake and risk of postmenopausal breast cancer in the NIH-AARP Diet and Health Study"

Supplementary table 1. Baseline characteristics by nutrient-adjusted ultra-processed food intake^a^ in NIH-AARP breast cancer analytic cohort (N=181,460)

|  | Q1 (N=36292) | Q2 (N=36292) | Q3 (N=36292) | Q4 (N=36292) | Q5 (N=36292) | Overall (N=181460) |
| --- | --- | --- | --- | --- | --- | --- |
| Median UPF intake, g/1000 kcal/day [IQR] | 146.8 (126.4, 162.5) | 206.1 (191.1, 222.7) | 284.6 (261.0, 311.8) | 417.3 (377.4, 466.6) | 756.0 (617.6, 1026.9) | 284.6 (191.1, 466.6) |
| Median age at baseline, years (IQR) | 63.9 (59.2, 67.4) | 63.4 (58.8, 67.0) | 62.8 (58.2, 66.6) | 62.1 (57.7, 66.1) | 60.9 (56.7, 65.3) | 62.6 (58.1, 66.6) |
| Median UPF intake, % kcal/day [IQR] | 34.0 (28.0, 40.1) | 41.5 (35.7, 47.7) | 43.3 (37.4, 49.6) | 44.8 (38.7, 51.1) | 47.2 (40.0, 54.8) | 42.2 (35.3, 49.2) |
| Median UPF intake, % g/day [IQR] | 8.6 (6.5, 11.2) | 13.1 (10.5, 16.6) | 18.1 (14.6, 22.7) | 25.0 (20.4, 31.0) | 40.3 (31.5, 51.9) | 18.4 (11.6, 28.7) |
| Race/ethnicity |  |  |  |  |  |  |
| American Indian/Alaskan Native | 130 (0.4%) | 85 (0.2%) | 118 (0.3%) | 112 (0.3%) | 154 (0.4%) | 599 (0.3%) |
| Asian | 1038 (2.9%) | 373 (1.0%) | 294 (0.8%) | 190 (0.5%) | 123 (0.3%) | 2018 (1.1%) |
| Hispanic | 993 (2.7%) | 805 (2.2%) | 693 (1.9%) | 576 (1.6%) | 426 (1.2%) | 3493 (1.9%) |
| Non-Hispanic Black | 1846 (5.1%) | 1754 (4.8%) | 2055 (5.7%) | 2226 (6.1%) | 2283 (6.3%) | 10164 (5.6%) |
| Non-Hispanic White | 31542 (86.9%) | 32695 (90.1%) | 32615 (89.9%) | 32661 (90.0%) | 32773 (90.3%) | 162286 (89.4%) |
| Pacific Islander | 88 (0.2%) | 41 (0.1%) | 47 (0.1%) | 28 (0.1%) | 22 (0.1%) | 226 (0.1%) |
| Unknown | 655 (1.8%) | 539 (1.5%) | 470 (1.3%) | 499 (1.4%) | 511 (1.4%) | 2674 (1.5%) |
| Smoking status |  |  |  |  |  |  |
| Never smoker | 16309 (44.9%) | 16576 (45.7%) | 16590 (45.7%) | 16336 (45.0%) | 14817 (40.8%) | 80628 (44.4%) |
| Former smoker | 13328 (36.7%) | 14108 (38.9%) | 14327 (39.5%) | 14074 (38.8%) | 14032 (38.7%) | 69869 (38.5%) |
| Current smoker | 5462 (15.1%) | 4546 (12.5%) | 4320 (11.9%) | 4832 (13.3%) | 6339 (17.5%) | 25499 (14.1%) |
| Unknown | 1193 (3.3%) | 1062 (2.9%) | 1055 (2.9%) | 1050 (2.9%) | 1104 (3.0%) | 5464 (3.0%) |
| Education level |  |  |  |  |  |  |
| Less than 12 years | 2200 (6.1%) | 2011 (5.5%) | 2204 (6.1%) | 2253 (6.2%) | 2664 (7.3%) | 11332 (6.2%) |
| 12 years, completed high school, GED | 8344 (23.0%) | 8911 (24.6%) | 9377 (25.8%) | 10047 (27.7%) | 10122 (27.9%) | 46801 (25.8%) |
| Post-high school training | 3570 (9.8%) | 3681 (10.1%) | 3918 (10.8%) | 4084 (11.3%) | 4244 (11.7%) | 19497 (10.7%) |
| Some college | 9118 (25.1%) | 9084 (25.0%) | 8887 (24.5%) | 8805 (24.3%) | 8882 (24.5%) | 44776 (24.7%) |
| College graduate | 11669 (32.2%) | 11504 (31.7%) | 10783 (29.7%) | 10021 (27.6%) | 9151 (25.2%) | 53128 (29.3%) |
| Unknown | 1391 (3.8%) | 1101 (3.0%) | 1123 (3.1%) | 1082 (3.0%) | 1229 (3.4%) | 5926 (3.3%) |
| Physical activity level |  |  |  |  |  |  |
| Never/rarely | 7402 (20.4%) | 7320 (20.2%) | 7604 (21.0%) | 8366 (23.1%) | 9910 (27.3%) | 40602 (22.4%) |
| Low (1-3 times a month) | 4593 (12.7%) | 4909 (13.5%) | 5045 (13.9%) | 5468 (15.1%) | 5789 (16.0%) | 25804 (14.2%) |
| Moderate (1-2 times per week) | 7148 (19.7%) | 7609 (21.0%) | 7859 (21.7%) | 7926 (21.8%) | 7390 (20.4%) | 37932 (20.9%) |
| High (≥3 times a week) | 16582 (45.7%) | 16011 (44.1%) | 15387 (42.4%) | 14085 (38.8%) | 12770 (35.2%) | 74835 (41.2%) |
| Unknown | 567 (1.6%) | 443 (1.2%) | 397 (1.1%) | 447 (1.2%) | 433 (1.2%) | 2287 (1.3%) |
| Alcohol intake |  |  |  |  |  |  |
| 0 drinks per day | 10322 (28.4%) | 9547 (26.3%) | 9982 (27.5%) | 10876 (30.0%) | 13074 (36.0%) | 53801 (29.6%) |
| <1 drink equivalents per day | 19661 (54.2%) | 21626 (59.6%) | 21992 (60.6%) | 21452 (59.1%) | 19604 (54.0%) | 104335 (57.5%) |
| 1-2 drink equivalents per day | 4837 (13.3%) | 4217 (11.6%) | 3568 (9.8%) | 3244 (8.9%) | 2622 (7.2%) | 18488 (10.2%) |
| 3-4 drink equivalents per day | 870 (2.4%) | 603 (1.7%) | 460 (1.3%) | 457 (1.3%) | 626 (1.7%) | 3016 (1.7%) |
| ≥5 drink equivalents per day | 602 (1.7%) | 299 (0.8%) | 290 (0.8%) | 263 (0.7%) | 366 (1.0%) | 1820 (1.0%) |
| BMI category |  |  |  |  |  |  |
| <18.5 kg/m^2^ | 744 (2.1%) | 591 (1.6%) | 543 (1.5%) | 439 (1.2%) | 435 (1.2%) | 2752 (1.5%) |
| 18.5 through <25 kg/m^2^ | 18323 (50.5%) | 16830 (46.4%) | 15129 (41.7%) | 13336 (36.7%) | 11314 (31.2%) | 74932 (41.3%) |
| 25 through <30 kg/m^2^ | 10249 (28.2%) | 11130 (30.7%) | 11679 (32.2%) | 12190 (33.6%) | 12182 (33.6%) | 57430 (31.6%) |
| ≥30 kg/m^2^ | 5575 (15.4%) | 6621 (18.2%) | 7905 (21.8%) | 9284 (25.6%) | 11288 (31.1%) | 40673 (22.4%) |
| Unknown | 1401 (3.9%) | 1120 (3.1%) | 1036 (2.9%) | 1043 (2.9%) | 1073 (3.0%) | 5673 (3.1%) |
| Marital status |  |  |  |  |  |  |
| Married | 15195 (41.9%) | 16014 (44.1%) | 16302 (44.9%) | 16759 (46.2%) | 15821 (43.6%) | 80091 (44.1%) |
| Widowed | 8702 (24.0%) | 8462 (23.3%) | 8299 (22.9%) | 8225 (22.7%) | 8082 (22.3%) | 41770 (23.0%) |
| Divorced or separated | 9419 (26.0%) | 9092 (25.1%) | 9012 (24.8%) | 8817 (24.3%) | 9774 (26.9%) | 46114 (25.4%) |
| Never married | 2536 (7.0%) | 2407 (6.6%) | 2367 (6.5%) | 2206 (6.1%) | 2285 (6.3%) | 11801 (6.5%) |
| Unknown | 440 (1.2%) | 317 (0.9%) | 312 (0.9%) | 285 (0.8%) | 330 (0.9%) | 1684 (0.9%) |
| Hormone replacement therapy duration |  |  |  |  |  |  |
| Never | 16760 (46.2%) | 15512 (42.7%) | 15487 (42.7%) | 15712 (43.3%) | 16190 (44.6%) | 79661 (43.9%) |
| < 5 yrs | 6313 (17.4%) | 6923 (19.1%) | 7101 (19.6%) | 7147 (19.7%) | 7250 (20.0%) | 34734 (19.1%) |
| 5-9 yrs | 4650 (12.8%) | 4966 (13.7%) | 5162 (14.2%) | 4997 (13.8%) | 4926 (13.6%) | 24701 (13.6%) |
| ≥10 yrs | 7823 (21.6%) | 8220 (22.6%) | 7812 (21.5%) | 7747 (21.3%) | 7170 (19.8%) | 38772 (21.4%) |
| Unknown | 746 (2.1%) | 671 (1.8%) | 730 (2.0%) | 689 (1.9%) | 756 (2.1%) | 3592 (2.0%) |
| Age at first period |  |  |  |  |  |  |
| ≤10 years | 2332 (6.4%) | 2177 (6.0%) | 2327 (6.4%) | 2516 (6.9%) | 2897 (8.0%) | 12249 (6.8%) |
| 11-12 years | 14449 (39.8%) | 15144 (41.7%) | 15328 (42.2%) | 15518 (42.8%) | 15682 (43.2%) | 76121 (41.9%) |
| 13-14 years | 15543 (42.8%) | 15471 (42.6%) | 15185 (41.8%) | 14886 (41.0%) | 14306 (39.4%) | 75391 (41.5%) |
| ≥15 years | 3805 (10.5%) | 3368 (9.3%) | 3344 (9.2%) | 3248 (8.9%) | 3282 (9.0%) | 17047 (9.4%) |
| Unknown | 163 (0.4%) | 132 (0.4%) | 108 (0.3%) | 124 (0.3%) | 125 (0.3%) | 652 (0.4%) |
| Oral contraceptive use |  |  |  |  |  |  |
| Never or < 1 yr | 23372 (64.4%) | 22625 (62.3%) | 22043 (60.7%) | 21356 (58.8%) | 20676 (57.0%) | 110072 (60.7%) |
| 1-4 yrs | 5710 (15.7%) | 5920 (16.3%) | 6179 (17.0%) | 6443 (17.8%) | 6766 (18.6%) | 31018 (17.1%) |
| ≥5 yrs | 6815 (18.8%) | 7451 (20.5%) | 7790 (21.5%) | 8209 (22.6%) | 8563 (23.6%) | 38828 (21.4%) |
| Unknown | 395 (1.1%) | 296 (0.8%) | 280 (0.8%) | 284 (0.8%) | 287 (0.8%) | 1542 (0.8%) |
| Parity |  |  |  |  |  |  |
| None | 5908 (16.3%) | 5563 (15.3%) | 5354 (14.8%) | 4993 (13.8%) | 5103 (14.1%) | 26921 (14.8%) |
| 1 | 3845 (10.6%) | 3618 (10.0%) | 3621 (10.0%) | 3652 (10.1%) | 3892 (10.7%) | 18628 (10.3%) |
| ≥2 | 26340 (72.6%) | 26950 (74.3%) | 27161 (74.8%) | 27495 (75.8%) | 27128 (74.7%) | 135074 (74.4%) |
| Unknown | 199 (0.5%) | 161 (0.4%) | 156 (0.4%) | 152 (0.4%) | 169 (0.5%) | 837 (0.5%) |
| First degree relative with breast cancer |  |  |  |  |  |  |
| No | 30072 (82.9%) | 30174 (83.1%) | 30119 (83.0%) | 30224 (83.3%) | 30284 (83.4%) | 150873 (83.1%) |
| Yes | 4491 (12.4%) | 4557 (12.6%) | 4606 (12.7%) | 4470 (12.3%) | 4303 (11.9%) | 22427 (12.4%) |
| Unknown | 1729 (4.8%) | 1561 (4.3%) | 1567 (4.3%) | 1598 (4.4%) | 1705 (4.7%) | 8160 (4.5%) |

Abbreviations: UPF, Ultra-processed food; HEI-2015, Healthy Eating Index-2015; IQR, interquartile range; BMI, body mass index

^a^ Quintiles of energy density-adjusted gram weight UPF (i.e., Nova group 4) intake (g/1000 kcal/day)

^b^ Column percentages might not add up to exactly 100% due to rounding.

Supplementary table 2. Dietary calcium, saturated fat, vegetable intake and meat intake (%) by Nova group in the NIH-AARP Diet and Health Study (N=181,460)

| Nutrient/Food group (%), mean (SD) | Nova 1 | Nova 2 | Nova 3 | Nova 4 |
| --- | --- | --- | --- | --- |
| Dietary Calcium | 56.4 (15.4) | 1.67 (3.0) | 9.75 (6.8) | 32.2 (12.8) |
| Saturated fat | 31.7 (11.9) | 12.3 (12.9) | 10.1 (6.2) | 45.9 (13.4) |
| Vegetables | 82.0 (9.7) | 0.0 (0.1) | 10.5 (7.2) | 7.55 (6.21) |
| Red meat | 98.6 (5.4) | 0 (0) | 0 (0) | 1.1 (1.7) |
| Processed meat | 0 (0) | 0 (0) | 27.2 (18.9) | 72.5 (19.3) |
| Total meat | 73.2 (13.9) | 0 (0) | 32.6 (38.0) | 15.2 (11.5) |

Nova 1=Unprocessed or minimally processed foods; Nova 2=Processed culinary ingredients; Nova 3=Processed foods; Nova 4=Ultra-processed foods. Abbreviation: SD = standard deviation.

Supplementary table 3. Association of quintiles of ultra-processed food intake (% kcal/day) with breast cancer risk in the NIH-AARP Diet and Health Study (N=181,460)

| Cancer type | N | Q1 HR (95% CI)  (N=36292) | Q2 HR (95% CI)  (N=36292) | Q3 HR (95% CI)  (N=36292) | Q4 HR (95% CI)  (N=36292) | Q5 HR (95% CI)  (N=36292) | *P* for trend^a^ |
| --- | --- | --- | --- | --- | --- | --- | --- |
| Breast cancer |  |  |  |  |  |  |  |
| No. of cases | 14484 | 2821 | 2967 | 3072 | 2817 | 2807 |  |
| Minimally adjusted model ^b^ |  | 1.00 (ref) | 1.06 (1.00-1.11) | 1.09 (1.03-1.15) | 1.00 (0.95-1.05) | 1.01 (0.96-1.06) | .63 |
| Multivariable adjusted model ^c^ |  | 1.00 (ref) | 1.04 (0.99-1.10) | 1.07 (1.01-1.12) | 0.99 (0.94-1.04) | 1.01 (0.96-1.06) | .62 |
| Multivariable adjusted + BMI model ^d^ |  | 1.00 (ref) | 1.04 (0.98-1.09) | 1.06 (1.01-1.11) | 0.98 (0.93-1.03) | 1.00 (0.95-1.06) | .40 |
| ER positive breast cancer |  |  |  |  |  |  |  |
| No. of cases | 8366 | 1613 | 1687 | 1792 | 1703 | 1571 |  |
| Minimally adjusted model ^b^ |  | 1.00 (ref) | 1.05 (0.98-1.13) | 1.11 (1.04-1.19) | 1.06 (0.99-1.14) | 1.00 (0.93-1.07) | .40 |
| Multivariable adjusted model ^c^ |  | 1.00 (ref) | 1.04 (0.97-1.12) | 1.09 (1.02-1.17) | 1.05 (0.98-1.13) | 1.01 (0.94-1.09) | .84 |
| Multivariable adjusted + BMI model ^d^ |  | 1.00 (ref) | 1.03 (0.96-1.10) | 1.08 (1.01-1.16) | 1.04 (0.97-1.11) | 1.00 (0.93-1.07) | .24 |
| ER negative breast cancer |  |  |  |  |  |  |  |
| No. of cases | 1579 | 308 | 329 | 344 | 288 | 310 |  |
| Minimally adjusted model ^b^ |  | 1.00 (ref) | 1.07 (0.92-1.26) | 1.12 (0.96-1.3) | 0.94 (0.8-1.10) | 1.02 (0.87-1.20) | .72 |
| Multivariable adjusted model ^c^ |  | 1.00 (ref) | 1.06 (0.91-1.24) | 1.10 (0.94-1.28) | 0.92 (0.78-1.08) | 1.01 (0.86-1.19) | .58 |
| Multivariable adjusted + BMI model ^d^ |  | 1.00 (ref) | 1.07 (0.91-1.25) | 1.10 (0.94-1.29) | 0.93 (0.79-1.09) | 1.01 (0.86-1.19) | .61 |
| Invasive |  |  |  |  |  |  |  |
| No. of cases | 11854 | 2319 | 2441 | 2517 | 2290 | 2287 |  |
| Minimally adjusted model ^b^ |  | 1.00 (ref) | 1.06 (1.00-1.12) | 1.09 (1.03-1.15) | 0.99 (0.93-1.05) | 1.00 (0.95-1.06) | .41 |
| Multivariable adjusted model ^c^ |  | 1.00 (ref) | 1.04 (0.99-1.10) | 1.06 (1.00-1.12) | 0.97 (0.92-1.03) | 1.00 (0.94-1.06) | .33 |
| Multivariable adjusted + BMI model ^d^ |  | 1.00 (ref) | 1.04 (0.98-1.10) | 1.05 (0.99-1.11) | 0.96 (0.91-1.02) | 0.99 (0.93-1.05) | .19 |
| Ductal carcinoma in situ |  |  |  |  |  |  |  |
| No. of cases | 2287 | 420 | 459 | 483 | 469 | 456 |  |
| Minimally adjusted model ^b^ |  | 1.00 (ref) | 1.10 (0.96-1.25) | 1.15 (1.01-1.31) | 1.12 (0.98-1.27) | 1.10 (0.96-1.25) | .20 |
| Multivariable adjusted model ^c^ |  | 1.00 (ref) | 1.09 (0.96-1.25) | 1.14 (1.00-1.31) | 1.12 (0.98-1.28) | 1.12 (0.98-1.28) | .12 |
| Multivariable adjusted + BMI model ^d^ |  | 1.00 (ref) | 1.09 (0.95-1.24) | 1.14 (1.00-1.30) | 1.11 (0.97-1.27) | 1.11 (0.97-1.28) | .13 |

Abbreviations: HR, hazard ratio; CI, confidence interval; BMI, body mass index

^a^ Each quintile was assigned to its median value and treated as a continuous variable.

^b^ Adjusted for age in years (underlying time metric) and total daily energy (kcal/day) for nutrient density adjustment.

^c^ Adjusted for age in years (underlying time metric), total daily energy (kcal/day), race/ethnicity (American Indian/Alaskan Native, Asian, Hispanic, Non-Hispanic Black, Non-Hispanic White, Pacific Islander, Unknown), smoking by intensity (cigarettes per day: 1-10, 11-20, 21-30, 31-40, 41-60, 61+) and time since cessation (10+ years ago, 5-9 years ago, 1-4 years ago, within the last year), education level (<12 years; 12 years, completed high school, or GED; post-high school training; some college; college and post graduate; unknown), physical activity level (never/rarely, 1-3 times/month, 1-2 times/week, ≥3 times/week, unknown), alcohol intake (0 drinks/day, < 1 drink/day, 1-2 drinks/day, 3-4 drinks/day, ≥5 drinks/day), first degree relative with breast cancer (yes/no), marital status (married, widowed, divorced or separated, never married, unknown), duration of HRT (never, <5, 5-9, ≥10 years, unknown), age at first menstrual period (<11, 11-12, 13-14, ≥15 years old, unknown), duration of oral contraceptive use (never or <1, 1-4, ≥5 years, unknown), and parity (0, 1, ≥2, unknown).

^d^ Multivariable model further adjusted for body mass index category (<18.5 kg/m2, 18.5 to <25 kg/m2, 25 to <30 kg/m2, ≥30 kg/m2, unknown).

Supplementary table 4. Association of quintiles of ultra-processed food intake (%g/day) with breast cancer risk in the NIH-AARP Diet and Health Study (N=181,460)

| Cancer type | N | Q1 HR (95% CI)  (N=36292) | Q2 HR (95% CI)  (N=36292) | Q3 HR (95% CI)  (N=36292) | Q4 HR (95% CI)  (N=36292) | Q5 HR (95% CI)  (N=36292) | *P*-trend^a^ |
| --- | --- | --- | --- | --- | --- | --- | --- |
| Breast cancer |  |  |  |  |  |  |  |
| No. of cases | 14484 | 2891 | 2961 | 2973 | 2919 | 2740 |  |
| Minimally adjusted model ^b^ |  | 1.00 (ref) | 1.01 (0.96-1.07) | 1.02 (0.97-1.08) | 1.01 (0.96-1.06) | 0.96 (0.91-1.02) | .08 |
| Multivariable adjusted model ^c^ |  | 1.00 (ref) | 1.01 (0.96-1.06) | 1.02 (0.97-1.08) | 1.02 (0.97-1.07) | 0.99 (0.93-1.04) | .52 |
| Multivariable adjusted + BMI model ^d^ |  | 1.00 (ref) | 1.00 (0.95-1.05) | 1.01 (0.96-1.07) | 1.00 (0.95-1.05) | 0.95 (0.90-1.01) | .06 |
| ER positive breast cancer |  |  |  |  |  |  |  |
| No. of cases | 8366 | 1677 | 1729 | 1728 | 1691 | 1541 |  |
| Minimally adjusted model ^b^ |  | 1.00 (ref) | 1.03 (0.96-1.10) | 1.04 (0.97-1.12) | 1.04 (0.97-1.11) | 0.98 (0.92-1.05) | .40 |
| Multivariable adjusted model ^c^ |  | 1.00 (ref) | 1.02 (0.95-1.09) | 1.04 (0.97-1.12) | 1.05 (0.98-1.12) | 1.01 (0.94-1.08) | .84 |
| Multivariable adjusted + BMI model ^d^ |  | 1.00 (ref) | 1.01 (0.94-1.08) | 1.02 (0.95-1.09) | 1.01 (0.95-1.09) | 0.96 (0.90-1.04) | .24 |
| ER negative breast cancer |  |  |  |  |  |  |  |
| No. of cases | 1579 | 301 | 334 | 312 | 316 | 316 |  |
| Minimally adjusted model ^b^ |  | 1.00 (ref) | 1.10 (0.94-1.29) | 1.03 (0.88-1.21) | 1.06 (0.90-1.24) | 1.09 (0.92-1.27) | .52 |
| Multivariable adjusted model ^c^ |  | 1.00 (ref) | 1.09 (0.93-1.27) | 1.01 (0.86-1.19) | 1.02 (0.87-1.20) | 1.05 (0.90-1.24) | .79 |
| Multivariable adjusted + BMI model ^d^ |  | 1.00 (ref) | 1.09 (0.93-1.27) | 1.02 (0.87-1.20) | 1.03 (0.88-1.21) | 1.07 (0.91-1.26) | .66 |
| Invasive |  |  |  |  |  |  |  |
| No. of cases | 11854 | 2374 | 2428 | 2459 | 2401 | 2192 |  |
| Minimally adjusted model ^b^ |  | 1.00 (ref) | 1.01 (0.96-1.07) | 1.03 (0.97-1.09) | 1.01 (0.96-1.07) | 0.94 (0.89-1.00) | .02 |
| Multivariable adjusted model ^c^ |  | 1.00 (ref) | 1.01 (0.95-1.07) | 1.04 (0.98-1.10) | 1.03 (0.97-1.09) | 0.97 (0.91-1.03) | .20 |
| Multivariable adjusted + BMI model ^d^ |  | 1.00 (ref) | 1.00 (0.95-1.06) | 1.02 (0.97-1.08) | 1.00 (0.95-1.06) | 0.93 (0.88-0.99) | .01 |
| Ductal carcinoma in situ |  |  |  |  |  |  |  |
| No. of cases | 2287 | 453 | 453 | 450 | 452 | 479 |  |
| Minimally adjusted model ^b^ |  | 1.00 (ref) | 0.99 (0.87-1.12) | 0.98 (0.86-1.12) | 0.99 (0.86-1.12) | 1.06 (0.93-1.20) | .32 |
| Multivariable adjusted model ^c^ |  | 1.00 (ref) | 0.97 (0.85-1.11) | 0.97 (0.85-1.10) | 0.98 (0.86-1.12) | 1.07 (0.94-1.22) | .17 |
| Multivariable adjusted + BMI model ^d^ |  | 1.00 (ref) | 0.97 (0.85-1.10) | 0.96 (0.84-1.09) | 0.97 (0.85-1.11) | 1.06 (0.92-1.21) | .26 |

Abbreviations: HR, hazard ratio; CI, confidence interval; BMI, body mass index

^a^ Each quintile was assigned to its median value and treated as a continuous variable.

^b^ Adjusted for age in years (underlying time metric) and total daily energy (kcal/day) for nutrient density adjustment.

^c^ Adjusted for age in years (underlying time metric), total daily energy (kcal/day), race/ethnicity (American Indian/Alaskan Native, Asian, Hispanic, Non-Hispanic Black, Non-Hispanic White, Pacific Islander, Unknown), smoking by intensity (cigarettes per day: 1-10, 11-20, 21-30, 31-40, 41-60, 61+) and time since cessation (10+ years ago, 5-9 years ago, 1-4 years ago, within the last year), education level (<12 years; 12 years, completed high school, or GED; post-high school training; some college; college and post graduate; unknown), physical activity level (never/rarely, 1-3 times/month, 1-2 times/week, ≥3 times/week, unknown), alcohol intake (0 drinks/day, < 1 drink/day, 1-2 drinks/day, 3-4 drinks/day, ≥5 drinks/day), first degree relative with breast cancer (yes/no), marital status (married, widowed, divorced or separated, never married, unknown), duration of HRT (never, <5, 5-9, ≥10 years, unknown), age at first menstrual period (<11, 11-12, 13-14, ≥15 years old, unknown), duration of oral contraceptive use (never or <1, 1-4, ≥5 years, unknown), and parity (0, 1, ≥2, unknown).

^d^ Multivariable model further adjusted for body mass index category (<18.5 kg/m2, 18.5 to <25 kg/m2, 25 to <30 kg/m2, ≥30 kg/m2, unknown).

Supplementary figure 1. Substitution analyses estimating the association for substituting intake of a select dietary factor from Nova group 4 (UPF) sources for Nova groups 1-3 (non-UPF) sources and addition analyses for the mutually adjusted associations for a select dietary factor from UPF and non-UPF sources

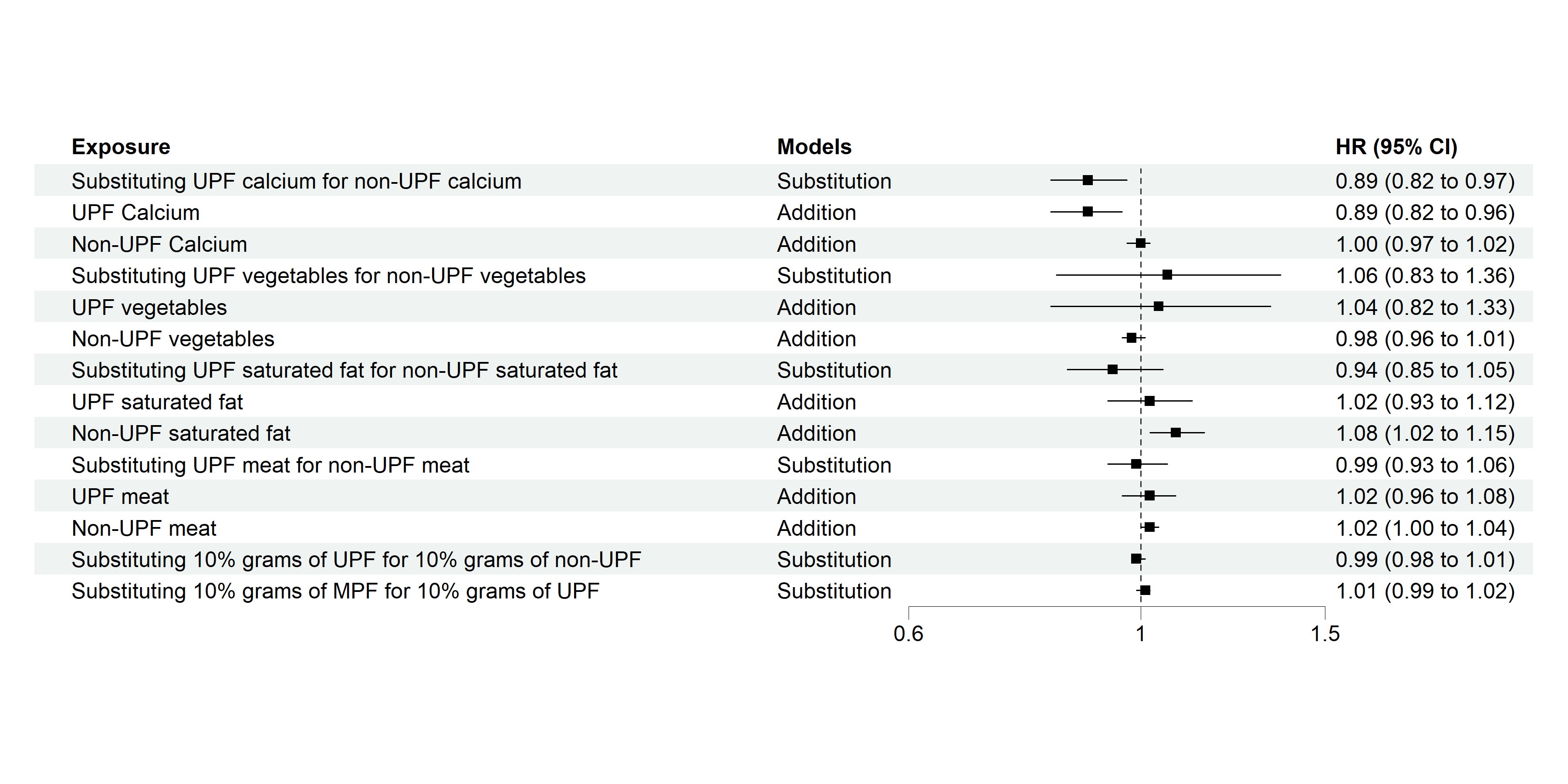

Abbreviations: UPF, ultra-processed food; HR, hazard ratio; CI, confidence interval; MPF, minimally processed food (Nova group 1)

Hazard ratios are estimated using Cox proportional hazard regression models with age as the underlying time metric adjusted total daily energy (kcal/day), race/ethnicity (American Indian/Alaskan Native, Asian, Hispanic, Non-Hispanic Black, Non-Hispanic White, Pacific Islander, Unknown), smoking by intensity (cigarettes per day: 1-10, 11-20, 21-30, 31-40, 41-60, >60) and time since cessation (≥10 years ago, 5-9 years ago, 1-4 years ago, within the last year), education level (11 years or less; 12 years, completed high school, or GED; post-high school training; some college; college and post graduate; unknown), physical activity level (never/rarely, low, moderate, high, unknown), alcohol intake (0 drinks/day, < 1 drink/day, 1-2 drinks/day, 3-4 drinks/day, ≥ 5 drinks/day), first degree relative with breast cancer (yes/no), marital status (married, widowed, divorced or separated, never married, unknown), duration of HRT (never, <5, 5-9, ≥10 years, unknown), age at first menstrual period (<11, 11-12, 13-14, ≥15 years old, unknown), duration of oral contraceptive use (never or <1, 1-4, ≥5 years, unknown), and parity (0, 1, ≥2, unknown). The hazard ratio for **calcium** is calculated per **300 mg/day increase**, for **saturated fat** per **10 g/day increase**, and for **vegetables and meat** per **serving per day.** In the **substitution models**, sources from the Nova group 4 were substituted for sources from Nova groups 1-3. The **addition model** accounts for mutual adjustments between sources from both Nova 1-3 and Nova 4 groups.
